# High-risk alcohol consumption may increase the risk of SARS-CoV-2 seroconversion: a prospective seroepidemiologic cohort study among American college students

**DOI:** 10.1101/2021.08.03.21261444

**Authors:** Sina Kianersi, Christina Ludema, Jonathan T. Macy, Chen Chen, Molly Rosenberg

## Abstract

**Aims:** To estimate the associations between high-risk alcohol consumption and (1) SARS-CoV-2 seroconversion, (2) self-reported new SARS-CoV-2 infection, and (3) symptomatic COVID-19.

**Design:** Prospective cohort

**Setting:** Indiana University Bloomington (IUB), a public university of 34,660 students in southern Indiana.

**Participants:** At the beginning of the fall 2020 semester, we randomly sampled N=1,267 IU undergraduate students, aged 18 years or older and residing in Monroe County, IN.

**Measurements:** Primary exposure was high-risk alcohol consumption measured with the AUDIT questionnaire. We used an AUDIT score of 8 or more as the cut-off score when detecting high-risk alcohol consumption. Primary outcome was SARS-CoV-2 seroconversion, assessed with two SARS-CoV-2 antibody tests, at baseline and endline. Secondary outcomes were a) self-reported new SARS-CoV-2 infection at the study endline, and b) self-reported symptomatic COVID-19 at baseline.

**Findings:** Prevalence of high-risk alcohol consumption was 34%. We found that students with high-risk alcohol consumption status had 2.34 [95% CI: (1.29, 4.24)] times the risk of SARS-CoV-2 seroconversion and 1.89 [95% CI: (1.08, 3.32)] times the risk of self-reporting a positive SARS-CoV-2 infection, compared to students with no such risk. Moreover, students with high-risk alcohol consumption were 18% more likely to develop symptomatic COVID-19, though this association was not statistically significant. Similar results were found after adjusting for sex at birth, race, and year in school. Findings from sensitivity analyses corroborated these results and suggested potential for a dose-response relationship.

**Conclusions:** In this sample of American college students, high-risk alcohol consumption was associated with higher risk for SARS-CoV-2 seroconversion/infection. These findings could have implications for colleges’ reopening planning in fall 2021.

## INTRODUCTION

### Background and rationale

Coronavirus disease 2019 (COVID-19), the disease caused by severe acute respiratory syndrome coronavirus 2 (SARS-CoV-2), has a major public health burden on college campuses. As of May 26, 2021, more than 700,000 SARS-CoV-2 infections have been reported from colleges and universities in the U.S., with the vast majority of the cases among students (1). COVID-19 has a wide range of symptoms, such as fever, cough, fatigue, and dyspnea (2). In some cases, COVID-19 causes long-lasting symptoms (long COVID (3)), such as loss of smell, impaired concentration, and memory problems among young adults (4). Acquiring COVID-19 and outbreaks of this disease on college campuses adversely impact students’ mental health and school performance and result in an increase in missed school days through isolation or quarantine requirements (5, 6). Finally, SARS-CoV-2 infection spread among college students can overflow into other segments of the community with higher risk for severe COVID-19 outcomes. Early increases in COVID-19 cases among college-age (18-24 years old) adults have been followed by increases in cases among older adults who are at higher risk of severe disease (7-9). Identifying modifiable risk factors for SARS-CoV-2 transmission among college students is imperative to prevent and control COVID-19 outbreaks on college campuses as well as among more vulnerable subpopulations in the community.

Alcohol consumption is an underexplored yet plausible risk factor for SARS-CoV-2 infection and transmission. It is a prevalent modifiable risky behavior, particularly among college students (10). In 2018, 51% of college-age adults reported drinking alcohol in the past 30 days, 24% reported binge drinking, and 6% reported heavy drinking (11). Alcohol consumption might increase individuals’ susceptibility to SARS-CoV-2 infection through two interrelated pathways, cognitive/behavioral and pathophysiological pathways.

Alcohol consumption causes cognitive distortion and brings about behavioral changes that could increase the risk of SARS-CoV-2 transmission and infection (12-14). It weakens vigilance, information processing, spatial working memory, and performance of complex tasks, and increases impulsivity (15-17). These cognitive changes plausibly disrupt compliance with COVID-19 protective behaviors, including mask wearing and physical distancing (12, 13, 18). Moreover, the alcohol use and cognitive distortion relationship might be cyclical. Young adults tend to drink more alcohol in groups (19), and more alcohol consumption exacerbates cognitive distortion which consequently can result in more non-compliance with COVID-19 protective measures (20-22).

Moreover, alcohol consumption impairs innate and adaptive immune subsystems’ responses to respiratory infections (23-26), through disrupting various immunological functionalities, such as reducing T-cell and B-cell count, impairing neutrophil production, and damaging alveolar barrier function (23-25). Similarly, we expect pathophysiological changes in the lungs due to alcohol consumption could contribute to SARS-CoV-2 infections (24). Lastly, alcohol consumption has also been found to increase susceptibility to respiratory complications, such as pneumonia (27) and acute respiratory distress syndrome (28). Hence, alcohol consumption might worsen COVID-19 prognosis as well.

The association between alcohol consumption and COVID-19 is not well understood. In commentaries and a non-quantitative review, researchers have suggested that the associations between alcohol use and COVID-19 incidence as well as COVID-19 severity need to be evaluated (24, 29-31). However, no quantitative study has been conducted to evaluate the associations between alcohol consumption and COVID-19 incidence and severity among college students, a population with prevalent excessive alcohol drinking and frequent COVID-19 outbreaks.

### Objectives

The primary objective was to longitudinally evaluate the association between high-risk alcohol consumption and SARS-CoV-2 seroconversion among college students. We hypothesized that students with high-risk alcohol consumption were more likely to experience SARS-CoV-2 seroconversion. Because seroconversion may have been imperfectly detected with our antibody tests, our secondary objective was to evaluate the association between high-risk alcohol consumption and self-reported positive SARS-CoV-2 PCR with reverse transcription (RT–PCR) testing history. We further assessed the association between high-risk alcohol consumption and symptomatic COVID-19 as another secondary outcome.

## METHODS

These results are presented using the STROBE (32) guidelines. The parent study was a randomized controlled trial evaluating the effect of receiving SARS-CoV-2 antibody test results on participants’ compliance with protective behavior against COVID-19 (33). Details about the parent study and overall study design are published elsewhere (14, 34).

### Study design

We used an observational, prospective exploratory cohort study design. Study period was from September to November 2020. Payments (up to $30) were made to participants to compensate them for their time. The IU Human Subjects and Institutional Review boards approved the study protocol (Protocol #2008293852). Participants provided informed consent through an online eConsent framework.

### Study setting, participants, and procedures

We conducted this study on the Indiana University Bloomington (IUB) campus. IUB has a total undergraduate population of ∼34,660. Many COVID-19 restrictions were in place during the data collection phase, including mask wearing, physical distancing, hybrid and remote classes, class spacing, contact tracing, mitigation testing, and quarantine and self-isolation mandates (https://www.iu.edu/covid/index.html). We acquired a random sample of IUB undergraduate students (n=7,499). Inclusion criteria were: 1) age ≥18 years, 2) IUB undergraduate student in fall 2020, and 3) residing in Monroe County, Indiana.

We sent study invitation emails with information about the study and links to an eligibility screening online survey to the 7,499 sampled students. Eligible students were directed to an online eConsent form with more information about the study. Students who consented to participate could schedule a baseline antibody testing appointment and complete the online baseline survey (34). This survey included questions about participant demographics, SARS-CoV-2 testing history, and alcohol use.

Between September 14-30, we conducted in-person SARS-CoV-2 baseline antibody tests on the IUB campus. We asked participants to reschedule their appointments if they were experiencing COVID-19 symptoms, had tested positive for SARS-CoV-2 in the last two weeks before their appointment, or had been directed to isolate or quarantine. Antibody testing results were entered into the REDCap (Research Electronic Data Capture (35, 36)) data capturing system (14).

Four follow-up online surveys were administered every two weeks after the baseline antibody test visit, starting September 28, 2020. In each follow-up survey, participants self-reported the quantity and frequency of their alcohol drinking over the last week. Using the same laboratory antibody testing procedures discussed above, we tested participants for SARS-CoV-2 antibodies at endliine. Lastly, on the fourth follow-up survey (endline survey), participants self-reported their RT-PCR SARS-CoV-2 testing history since baseline.

### Variables

#### Primary exposure

The main exposure was high-risk alcohol consumption, measured with Alcohol Use Disorders Identification Test (AUDIT) (self-report version) (37). Previous studies have established AUDIT as a valid measurement tool for use among young adults and college students (38, 39). AUDIT has ten questions. The first three are about frequency and quantity of alcohol consumption, questions four to six are about drinking behavior during the last year, and the last four questions are about drinking problems during the last year. Each question can contribute a score from 0 to 4, and correspondingly a total AUDIT score can range from 0 to 40 (37). In our main analysis, we used an AUDIT score of 8 or more (AUDIT≥8 vs. AUDIT<8) as the cut-off score for high-risk drinking, as established in prior studies (37, 39).

#### Secondary exposures for sensitivity analyses

We used three set of secondary exposures in our sensitivity analyses. To explore the sensitivity of our findings to choices around AUDIT score cut-offs we used other AUDIT cut-off points, from 3 to 13. Because the number of seroconversion outcomes was small (≤5) in one of the exposure groups for cut-offs below 3 and above 13, we did not explore these cut-offs. Another secondary exposure was AUDIT-C score. AUDIT-C is an effective and brief three-question measurement tool for detecting high-risk alcohol consumption (40), validated for use among college students (41). We used a cut-off score of 7 for males and 5 for females when using AUDIT-C to identify at-risk drinkers (41). Lastly, in each of the four follow-up surveys, we collected weekly alcohol use data using a quantity-frequency measure. The questions used for collecting these data were similar to that from the Behavioral Risk Factor Surveillance System (42), though slightly reworded. Using these data, we made the following secondary exposures: a) any drinking during the study follow-up, b) heavy drinking (>14 drinks per week for men and >7 drinks per week for women in any of the four follow-up surveys)

#### Primary outcome

The main outcome was SARS-CoV-2 seroconversion. For primary outcome analyses, we had an additional exclusion criterion. Participants needed to be seronegative for SARS-CoV-2 antibodies at baseline (n=1027). Seroconversion was defined as having a negative SARS-CoV-2 antibody test result at baseline and a positive one at endline. We used SARS-CoV-2 IgM/IgG rapid assay kit (Colloidal Gold method) to test participants for SARS-CoV-2 IgM and IgG antibodies. Compared to a CLIA Lab-based validation analysis, our BGI rapid kits showed a 64% of sensitivity and a 100% specificity. The antibody test result was interpreted as positive if one or both of IgG and IgM antibody types were detected in the blood sample.

#### Secondary outcomes

We chose two secondary outcomes:

##### 1) Self-reported new SARS-CoV-2 infections since baseline

participants self-reported their testing history for active SARS-CoV-2 infection in baseline and endline surveys. At baseline, they reported if they have ever been tested positive for SARS-CoV-2 infection. In endline survey, we asked participants if they have been tested for an active SARS-CoV-2 infection since baseline. Among those who responded “Yes” to this question, we asked about the results of their test. Only participants who self-reported a negative SARS-CoV-2 infection history at baseline and self-reported testing for SARS-CoV-2 active infection since baseline, in the endline survey, were eligible for this outcome analyses (n=518). New SARS-CoV-2 infection was defined as self-reporting a negative SARS-CoV-2 antibody test result in baseline and a positive one in endline surveys.

##### 2) Self-reports of symptomatic COVID-19

At baseline, among participants who self-reported that they have ever been tested positive for SARS-CoV-2 infection (n=128), we asked them to describe the symptoms of their SARS-CoV-2 infection. There were four response options for this question: “Asymptomatic”, “Mild”,” Moderate”,” Severe”. For this dichotomized secondary outcome, participants who reported “Mild”,” Moderate”, or “Severe” symptoms were recoded as symptomatic (Asymptomatic vs. Symptomatic).

#### Covariates

In the baseline survey, participants self-reported the following demographics: age (years), sex at birth (female vs. male), race (Asian, Black, multiracial, other, white), year in school (1^st^ - 4^th^ and 5^th^), residence (on-campus vs. off-campus), and Greek membership (yes vs. no). For analysis, we dichotomized age to distinguish between the typical range of college ages (18–22 years) and older. We also dichotomized the race variable (white vs. non-white). Participants had the option to choose “Don’t Know” when responding to the baseline survey questions. “Don’t know” responses were set to missing in the analysis.

### Study size

We did not perform power analysis for the current cohort study as this study was leveraged from the RCT study.

### Statistical methods

We used Poisson regression with a robust error variance (43) to estimate the unadjusted and adjusted risk ratios (RR) for the association between the primary exposure (high-risk alcohol consumption) and primary outcome (SARS-CoV-2 seroconversion) and secondary outcome (self-reported new SARS-CoV-2 infection). We used the same statistical models (43) to estimate the unadjusted and adjusted prevalence ratios (PR) for the associations between the primary exposure and symptomatic COVID-19. Poisson regression with a robust error variance is the method of choice when estimating PRs (44). The adjustment set in all multivariable models was sex at birth, dichotomized race, and year in school, measured in the baseline survey. We calculated the 95% confidence interval for all estimated RRs and PRs. In all models, we used complete case analysis.

In our sensitivity analysis, we used other AUDIT cut-offs (3-13), AUDIT-C, and quantity and frequency of alcohol consumption (any drinking and heavy drinking) and re-estimated the RR for the associations between these exposures and the outcomes. The data analysis was conducted using SAS software, Version 9.4 (Cary, NC, USA). We used Python for data visualization (version 3.7.6, Python Software Foundation, Beaverton, OR, US).

## RESULTS

### Participants

Of the 7,499 sampled IUB undergraduate students, 3,430 did not meet one or more of the inclusion criteria and 2,672 were non-responders (Figure 1). A total of 1,397 students consented to participate in the study (14). However, 130 of them did not complete any of the study procedures and 191 did not complete their baseline SARS-CoV-2 antibody testing. Moreover, 49 of the 1,076 participants who completed their baseline antibody test appointment tested positive at baseline and were excluded from the analysis regarding the primary outcome. Of the 1,027 participants who tested negative at baseline, n= 808 (79%) returned for their endline antibody test. A total of 736 participants completed all follow-up surveys.

**Figure 1.**
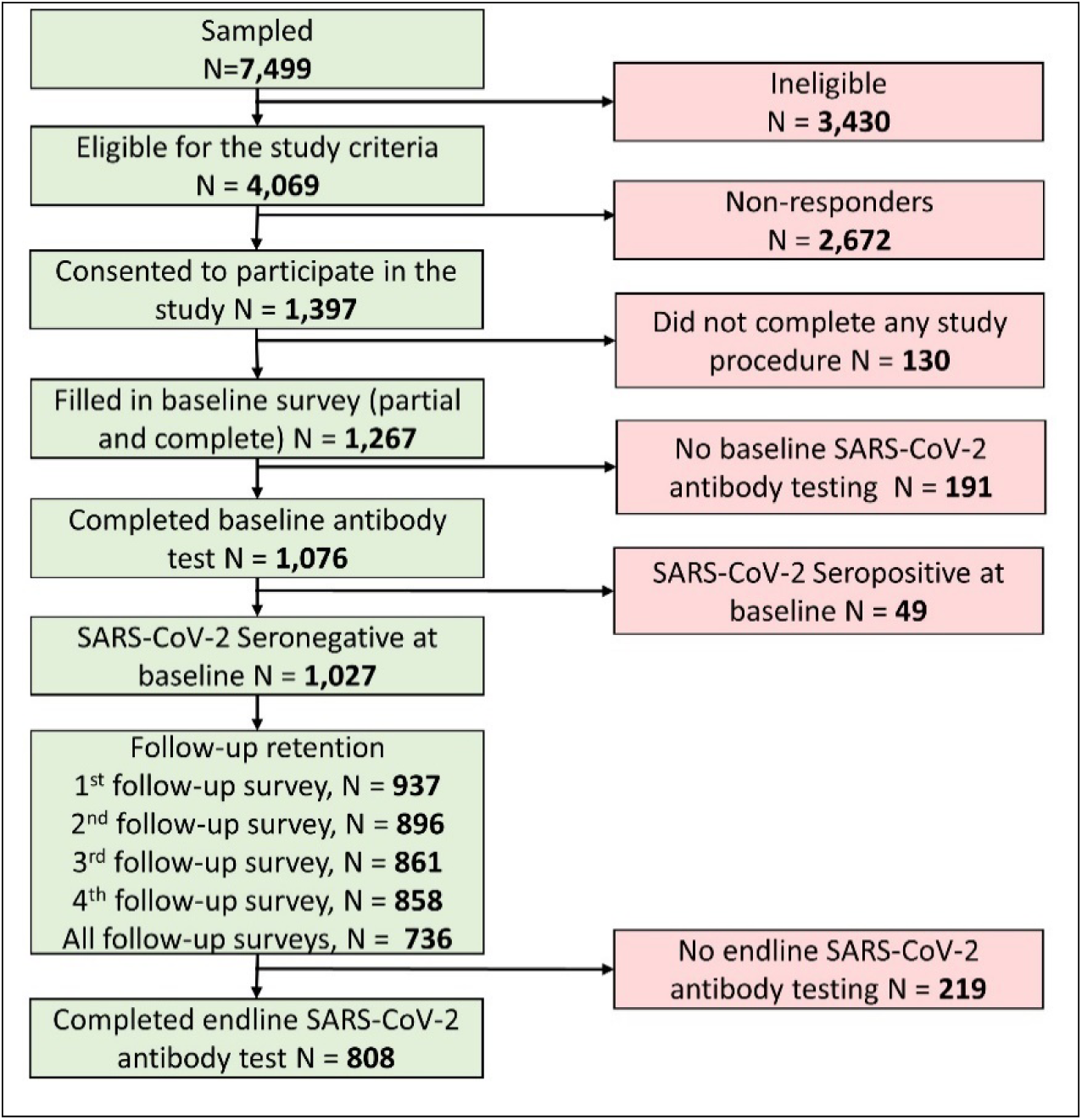
Flow diagram of the study sample.

### Descriptive data

Age median was 20 (interquartile range=2). Students were mostly female (63%), white (77%), senior undergraduate student (31%), and non-Greek affiliated (76%) (Table 1). Around 67% of participants reported living off-campus. There were significant sociodemographic differences between participants with high-risk alcohol consumption (AUDIT score ≥8) and low-risk alcohol consumption (AUDIT score <8). In chi-squared tests, sex at birth, race, year in school, residence, and Greek membership were associated with high-risk alcohol consumption. Participants with high-risk alcohol consumption status tended to be male, white, senior students, living off-campus, and affiliated with the Greek organizations. Study response rate was 29% (14). This response rate is above average and comparable to other similar studies (45-47). The retention rate at endline was 79%.

**Table 1.** Baseline characteristics of the study participants of 1,267 Indiana University undergraduate students, September 2020

|  | Total<br>N=1,267<br>n (%) | Primary exposure:<br>Total AUDIT Score<sup>a</sup> |  | p-value<sup>b</sup> |
| --- | --- | --- | --- | --- |
|  |  | AUDIT<8<br>n = 810 (66.5) | AUDIT≥8<br>n = 409 (33.6) |  |
| Socio-demographic characteristics |  |  |  |  |
| Age |  |  |  | 0.9100 |
| ≥22 years old | 116 (9.8) | 74 (9.7) | 38 (9.9) |  |
| <22 years old | 1071 (90.2) | 688 (90.3) | 345 (90.1) |  |
| Missing | 80 | 48 | 26 |  |
| Sex at birth |  |  |  | <.0001 |
| Female | 800 (63.4) | 563 (69.5) | 215 (52.7) |  |
| Male | 462 (36.6) | 247 (30.5) | 193 (47.3) |  |
| Missing | 5 | 0 | 1 |  |
| Race |  |  |  | <.0001<sup>c</sup> |
| Asian | 99 (7.8) | 82 (10.1) | 14 (3.4) |  |
| Black | 25 (2.0) | 20 (2.5) | 3 (0.7) |  |
| Multi-racial | 103 (8.2) | 74 (9.1) | 28 (6.9) |  |
| Other | 60 (4.8) | 44 (5.4) | 13 (3.2) |  |
| White | 975 (77.3) | 590 (72.8) | 351 (85.8) |  |
| Missing | 5 | 0 | 0 |  |
| Race dichotomized |  |  |  | <.0001 |
| White | 975 (77.3) | 590 (72.8) | 351 (85.8) |  |
| Non-White | 287 (22.7) | 220 (27.2) | 58 (14.2) |  |
| Missing | 5 | 0 | 0 |  |
| Year in school |  |  |  | <.0001 |
| 1<sup>st</sup> | 286 (22.7) | 204 (25.3) | 67 (16.4) |  |
| 2<sup>nd</sup> | 284 (22.5) | 191 (23.7) | 83 (20.3) |  |
| 3<sup>rd</sup> | 306 (24.3) | 201 (24.9) | 96 (23.5) |  |
| 4<sup>th</sup> and 5<sup>th</sup> | 384 (30.5) | 211 (26.2) | 163 (39.9) |  |
| Missing | 7 | 3 | 0 |  |
| Residence |  |  |  | 0.0001 |
| Off-campus | 850 (67.4) | 520 (64.3) | 307 (75.3) |  |
| On-campus | 411 (32.6) | 289 (35.7) | 101 (24.8) |  |
| Missing | 6 | 1 | 1 |  |
| Greek membership |  |  |  | <.0001 |
| No | 957 (76.0) | 656 (81.3) | 271 (66.3) |  |
| Yes | 303 (24.1) | 151 (18.7) | 138 (33.7) |  |
| Missing | 7 | 3 | 0 |  |
| Primary outcome |  |  |  |  |
| SARS-CoV-2 seroconversion, among those who were seronegative for SARS-CoV-2 at baseline (N=1027) |  |  |  | 0.0039 |
| Yes | 42 (5.2) | 20 (3.6) | 21 (8.5) |  |
| No | 766 (94.8) | 531 (96.4) | 226 (91.5) |  |
| Missing | 219 | 137 | 74 |  |
| Secondary outcomes |  |  |  |  |
| Self-reported new SARS-CoV-2 infections since baseline<sup>d</sup> |  |  |  | 0.0244 |
| Yes | 44 (8.6) | 23 (6.8) | 21 (12.8) |  |
| No | 465 (91.4) | 317 (93.2) | 143 (87.2) |  |
| Missing | 9 | 6 | 3 |  |

| Symptomatic COVID-19<sup>e</sup> |  |  |  | 0.0709 |
| --- | --- | --- | --- | --- |
| Yes | 95 (75.4) | 42 (68.9) | 48 (81.4) |  |
| No | 31 (24.6) | 19 (31.1) | 11 (18.6) |  |
| Missing | 2 | 2 | 0 |  |
| a. Overall, AUDIT score was missing for 48 participants<br>b. Chi-squared and Fisher's Exact tests were used to compare the groups<br>c. Fisher's Exact test due to small cell sizes<br>d. Among those who self-reported a negative SARS-CoV-2 infection history at baseline and self-reported testing for SARS-CoV-2 active infection since baseline, in the endline survey (n=518)<br>e. Among those who self-reported positive SARS-CoV-2 testing history at baseline (n=128) |  |  |  |  |

AUDIT score data were available for 1,219 participants (n-missing=48) (Table 1). AUDIT score median was 5 with an interquartile range of 7 (Supporting Information: Figure S1). Around 34% of participants were at high-risk alcohol consumption.

#### Primary outcome

Of the 808 participants who tested negative at baseline and completed the endline antibody test, 42 (5%) seroconverted; 21 (9%) of 247 participants with high-risk alcohol consumption status seroconverted while only 20 (4%) of 551 participants with low-risk alcohol consumption status seroconverted (Table 1, Figure 2). The proportion of missing values for primary outcome was similar across the low-risk and high-risk groups.

**Figure 2.**
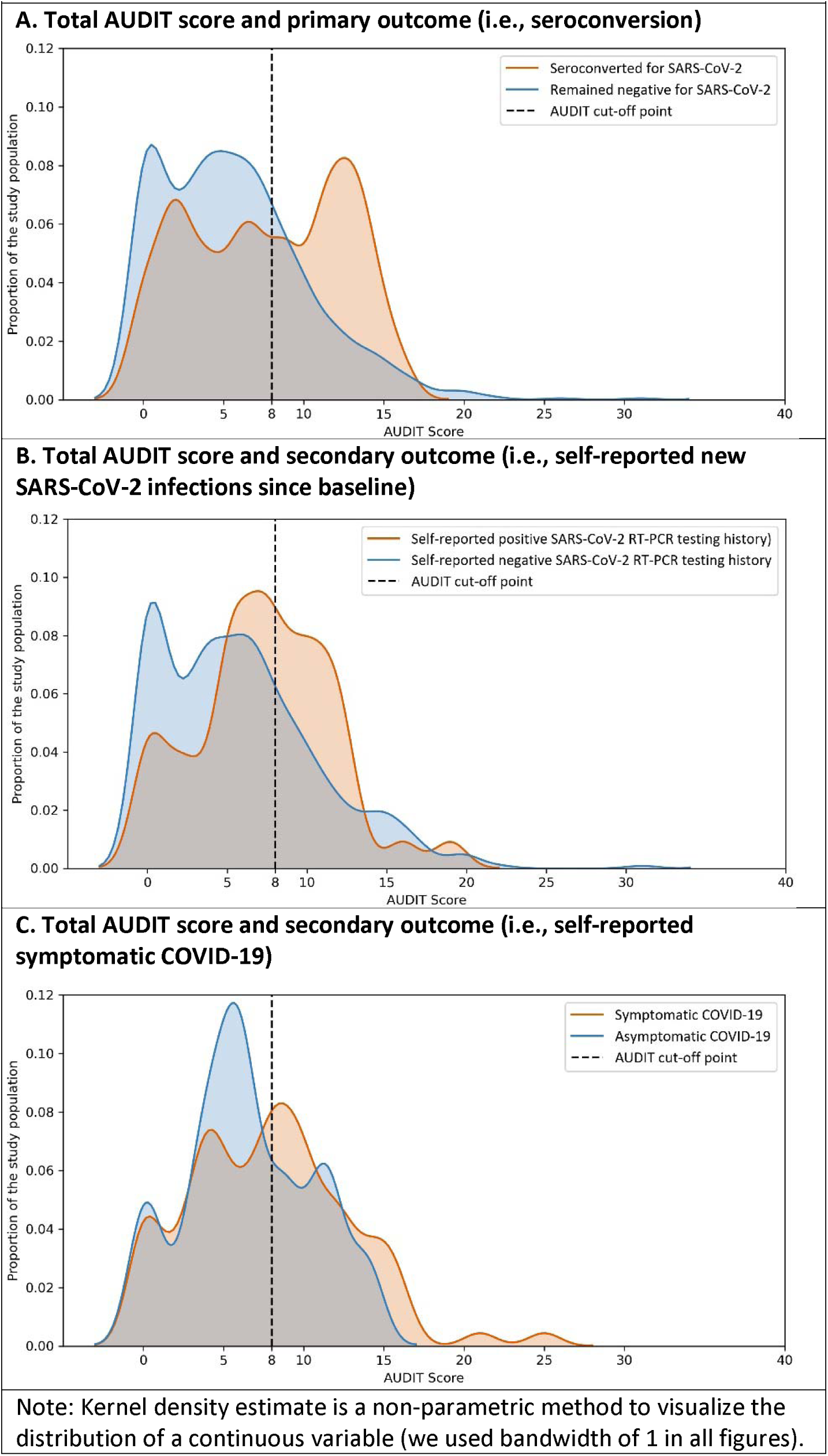
Kernel density estimates of AUDIT score by primary and secondary COVID-19 outcomes.

#### Secondary outcomes

Overall, 9% of participants who self-reported a negative SARS-CoV-2 infection history at baseline self-reported a positive SARS-CoV-2 infection in the endline survey. Moreover, among participants who self-reported that they have ever been tested positive for SARS-CoV-2 infection in the baseline survey (n=126), 75% reported experiencing symptomatic COVID-19.

### Main results

#### Primary exposure and primary/secondary outcomes

In crude analysis (Table 2), we found that students with high-risk alcohol consumption had 2.34 times the risk of SARS-CoV-2 seroconversion [RR (95% CI): 2.34 (1.29, 4.24)], and 1.89 times the risk of self-reporting a positive SARS-CoV-2 infection at endline [RR (95% CI): 1.89 (1.08, 3.32)], compared to students with low-risk alcohol consumption. At baseline, students with high-risk alcohol consumption status were 18% more likely to experience symptomatic COVID-19, compared to students with low-risk alcohol consumption, though this association was statistically insignificant [PR (95% CI): 1.18 (0.96, 1.46)]. Similar associations were found in multivariable models (Supporting Information: Table S1).

**Table 2.** Crude associations between alcohol consumption and COVID-19 outcome

|  |  |  |  |
| --- | --- | --- | --- |
| Primary Exposure | Primary outcome | Secondary outcomes |  |
|  | SARS-CoV-2 seroconversion at endline<br>Unadjusted RR | Self-reported new SARS-CoV-2 infections at endline<br>Unadjusted RR | Symptomatic COVID-19 self-report at baseline<br>Unadjusted PR |
| High-risk alcohol consumption assessed with AUDIT | n = 798 | n = 504 | n = 120 |
| Yes (AUDIT≥8) | **2.34 (1.29, 4.24)** | **1.89 (1.08, 3.32)** | 1.18 (0.96, 1.46) |
| No (AUDIT<8) | Ref. | Ref. | Ref. |
| Secondary Exposures |  |  |  |
| High-risk alcohol consumption assessed with AUDIT-C | n = 803 | n = 508 | n = 121 |
| Yes (AUDIT-C≥7 for males and AUDIT-C ≥5 for females) | **2.47 (1.35, 4.49)** | **2.45 (1.37, 4.37)** | 1.06 (0.86, 1.31) |
| No (AUDIT-C<7 for males and AUDIT-C <5 for females) | Ref. | Ref. | Ref. |
| Frequency and quantity of alcohol consumption |  |  |  |
| Any drinking | n = 768 | n = 493 | n = 86 |
| Yes | 1.31 (0.56, 3.06) | 1.98 (0.80, 4.91) | 0.87 (0.63, 1.21) |
| No | Ref. | Ref. | Ref. |
| Heavy drinking | n = 701 | n = 467 | n = 76 |
| Yes | **2.36 (1.23, 4.54)** | **2.51 (1.40, 4.50)** | 1.22 (0.94, 1.60) |
| No | Ref. | Ref. | Ref. |
| Boldface indicates significant value (p<0.05) |  |  |  |
| RR: Risk Ratio; PR: Prevalence Ratio |  |  |  |

#### Sensitivity analyses

##### Secondary exposures and primary/secondary outcomes

We found similar results when we used AUDIT-C instead of AUDIT as the exposure variable (Table 2, Supporting Information: Table S1). Moreover, we found that participants who reported heavy drinking in one or more of the follow-up surveys had 2.36 times the risk of SARS-CoV-2 seroconversion and 2.51 times the risk of self-reporting a new positive SARS-CoV-2 infection, compared to participants who did not report heavy drinking (Table 2). The magnitude of these associations increased when we controlled for the adjustment set (Supporting Information: Table S1). Lastly, the association between AUDIT score and SARS-CoV-2 seroconversion was generally low in magnitude with wide confidence intervals including the null when we used cut-offs below 6 (Figure 3). The association tended to increase when we used cut-offs of 6 or larger.

**Figure 3.**
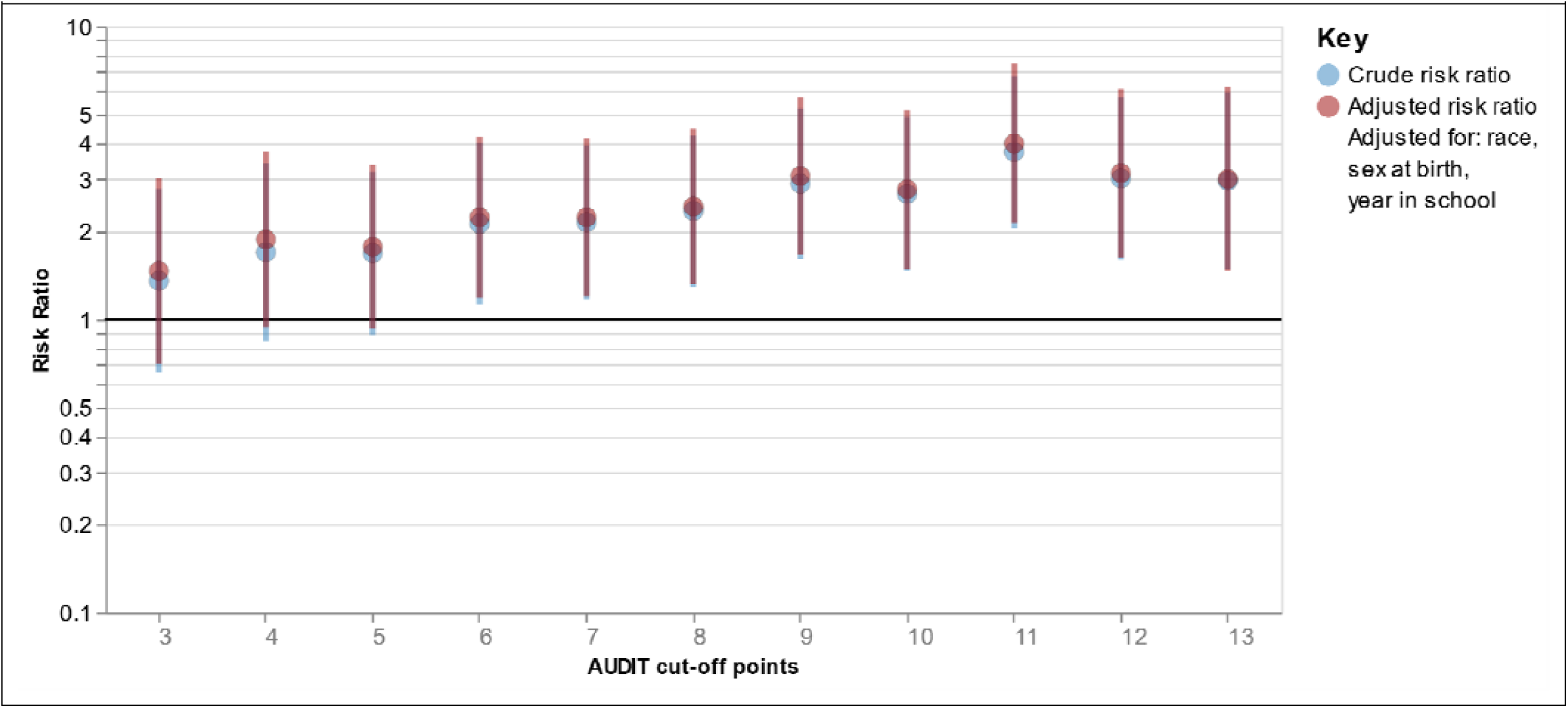
Association between AUDIT score dichotomized at different cut-off points and SARS-CoV-2 seroconversion.

## DISCUSSIONS

### Key results

We found that undergraduate students with high-risk alcohol consumption were at higher risk for SARS-CoV-2 seroconversion, compared to students with no such risk. We found similar results when we used an alternative outcome (self-reported new SARS-CoV-2 infection). Participants with high-risk alcohol consumption were more likely to experience symptomatic COVID-19, though this association was not statistically significant. In sensitivity analyses, we found similar results when we used other alcohol consumption exposures (other AUDIT cut-offs, AUDIT-C, and heavy drinking). The association between AUDIT score and seroconversion did not exist when using AUDIT score cut-offs below 6 but existed and seemingly increased for cut-offs above 6. This study highlights the important role alcohol may play in the spread of COVID-19 on college campuses.

### Interpretation

Few studies have evaluated similar associations between alcohol use and COVID-19. In our literature screening of over 660 titles on PubMed, we identified eight relevant study reports (48-55). Two studies did not find any association between alcohol consumption and COVID-19 (48, 51). Four studies identified excessive alcohol use as a risk factor for COVID-19 diagnosis (54), poor prognosis (53), and severity (49, 55). One study found excessive alcohol use as a risk factor for COVID-19 death in patients with obesity but not in those without obesity (52). One study found low-dose alcohol intake (<100g alcohol per week) a protective factor for COVID-19 hospitalization (50). Lastly, in a previous cross-sectional analysis report using baseline data of the RCT study, we found that drinking alcohol more than once a week increased the likelihood of SARS-CoV-2 seropositivity (14).

These studies were heterogenous in their methodology, target population, and exposure and outcome measurements. Only one study was conducted in the U.S. (among hospital patients) (54). No prior study was in college students or young adults, a general population among whom excessive alcohol drinking and SARS-CoV-2 infections are both prevalent. Previous studies measured alcohol consumption in different ways, such as quantity-frequency questionnaire (48-50), and semi-structured interviews (53). To our knowledge, no study used the validated AUDIT screening tool. AUDIT screens a longer period compared to other measurement tools, and it can identify harmful drinking pattern and chronic alcohol consumption which are linked to adverse physical consequences (56). Studies used different ways to measure COVID-19 outcome, such as RT-PCR test (52), electronic health records (54), or COVID-19 hospitalization (48, 50, 51). No study used SARS-CoV-2 seroconversion. Serological tests can detect previously infected individuals even if they were not tested for active infection using RT-PCR test.

We further observed that the magnitude of the association between dichotomized AUDIT score and SARS-CoV-2 seroconversion tended to increase as the AUDIT cut-offs increased from 6 to 13. These AUDIT scores correspond to hazardous alcohol consumption (37, 39). These findings suggest that there might be a dose-response relationship between alcohol consumption and SARS-CoV-2 seroconversion. Another study found that low-dose alcohol use is associated with lower risk for COVD-19 hospitalization (50). Previous studies have found similar protective associations for low to moderate alcohol use and other respiratory infections, such as the common cold (57). However, we did not find any protective relationship for low alcohol use against SARS-CoV-2 infection among our sample of college students.

### Strengths and limitations

#### Study design

Some aspects of our study design influence the interpretation of our findings. Our study design was observational and therefore inferring a causal relationship between alcohol use and SARS-CoV-2 seroconversion is difficult because of the potential for unmeasured confounding. Moreover, the associations between high-risk alcohol consumption and secondary outcome of symptomatic COVID-19 were evaluated cross-sectionally and with a small sample size. Thus, our ability to evaluate the temporal ordering between high-risk alcohol consumption and symptomatic COVID-19 outcome is limited. Nonetheless, we used a robuster study design (prospective cohort) with larger sample sizes when evaluating the associations between high-risk alcohol consumption and seroconversion and self-reported SARS-CoV-2 new infection outcomes.

#### Measures

We chose different measurement tools for assessing the exposure and outcome, each of which have some strengths and limitations. We used biological antibody testing to measure seroconversion outcome. Antibody testing kits can capture undetected previous SARS-CoV-2 infections. Yet, the antibody testing kits in this study had a low sensitivity. However, we used a secondary outcome (self-reported new SARS-CoV-2 infection) that does not depend on antibody positivity. Previously, we found a strong association between self-reported SARS-CoV-2 infection and antibody testing variables (14). Similarly, we used different measures to collect self-reported data on alcohol use, AUDIT (with different cut-offs), AUDIT-C, and quantity-frequency index. Because alcohol use data were self-reported, the data might suffer from recall and social desirability biases. However, all these measurement tools are validated. Collecting real-time alcohol use data using ecological momentary assessment tools might help to reduce these biases (58).

#### Generalizability

We used random sampling to identify our potential study participants. Because the demographics of IUB undergraduates are comparable to that of other large campuses, we might be able to generalize our findings to American college students. The response rates of 28.7% for the parent study might seem low. However, this would not weaken the generalizability of our finding. Our response rate is considered greater than average when compared to other studies on college campuses (46, 47).

## Conclusion

Our findings suggest that high-risk alcohol consumption is associated with higher risk for SARS-CoV-2 infection/seroconversion. These findings could have implications for colleges’ reopening planning in fall 2021. Even though effective interventions to reduce high-risk alcohol consumption might take more time to be tested and implemented at individual level, university policy makers can use our findings to predict locations in college towns where transmission might be more likely to occur (e.g., college town bars or Greek houses) and implement COVID-19 protective measures (e.g., face mask wearing) in such places, in fall 2021. More studies are needed to understand the extent of a causal relationship between alcohol consumption and SARS-CoV-2 infection.

## Supporting information

STROBE

## Data Availability

Data are available upon request.

## Funding

This study was supported by private contributions to the Indiana University Foundation. The United Arab Emirates provided the testing kits.

## Notes

### Competing Interest Statement

The authors have declared no competing interest.

### Clinical Trial

NCT04620798

### Author Declarations

The Indiana University Human Subjects and Institutional Review boards approved the study protocol (Protocol #2008293852).

