## Supporting Information: for "High-risk alcohol consumption may increase the risk of SARS-CoV-2 seroconversion: a prospective seroepidemiologic cohort study among American college students"

| Table S1. Adjusted associations between alcohol consumption and COVID-19 outcome | | | |
| --- | --- | --- | --- |
| Primary Exposure | Primary outcome | Secondary  outcomes | |
|  | SARS-CoV-2 seroconversion at endline | Self-reported new SARS-CoV-2 infections at endline | Symptomatic COVID-19 self-report at baseline^b^ |
|  | Unadjusted RR | Unadjusted RR | Unadjusted PR |
| High-risk alcohol consumption assessed with AUDIT | n = 797 | n = 503 | n = 119 |
| Yes (AUDIT≥8) | **2.43 (1.32, 4.46)** | **1.90 (1.08, 3.33)** | 1.18 (0.93, 1.49) |
| No (AUDIT<8) | Ref. | Ref. | Ref. |
| Secondary Exposures |  |  |  |
| High-risk alcohol consumption assessed with AUDIT-C | n = 801 | n = 506 | n = 121 |
| Yes (AUDIT-C≥7 for males and AUDIT-C ≥5 for females) | **2.61 (1.45, 4.72)** | **2.47 (1.36, 4.51)** | 0.99 (0.79, 1.23) |
| No (AUDIT-C<7 for males and AUDIT-C <5 for females) | Ref. | Ref. | Ref. |
| Frequency and quantity of alcohol consumption |  |  |  |
| Any drinking | n = 765 | n = 492 | n = 85 |
| Yes | 1.45 (0.61, 3.42) | 2.18 (0.89, 5.35) | 0.80 (0.56, 1.13) |
| No | Ref. | Ref. | Ref. |
| Heavy drinking | n = 699 | n = 466 | n = 76 |
| Yes | **2.71 (1.43, 5.15)** | **2.60 (1.42, 4.75)** | 1.27 (0.95, 1.70) |
| No | Ref. | Ref. | Ref. |
| All models in this tables were adjusted for sex at birth, race, year in school  Boldface indicates significant value (p<0.05)  RR: Risk Ratio; PR: Prevalence Ratio | | | |

Figure S1. AUDIT score distribution in the sample

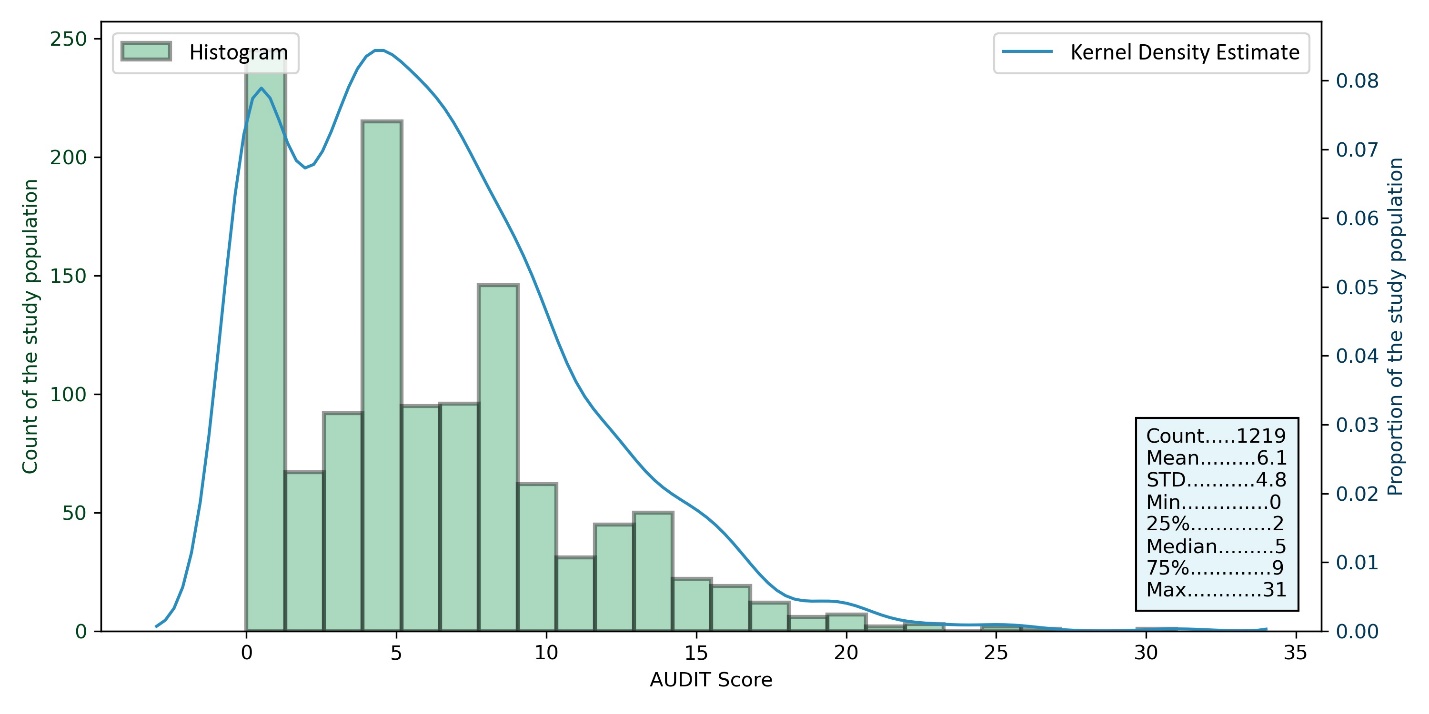
